# Proteomic analysis of human serum Extracellular Vesicles reveals early diagnostic markers for Amyotrophic Lateral Sclerosis

**DOI:** 10.1101/2023.07.26.23292854

**Authors:** Natasha Vassileff, Pascal Leblanc, Emilien Bernard, Anthony Fourier, Isabelle Quadrio, Rohan G. T. Lowe, Jereme G. Spiers, Andrew F. Hill, Lesley Cheng

## Abstract

Amyotrophic Lateral Sclerosis (ALS) is a neurodegenerative disease characterized by the deposition of misfolded proteins leading to the death of motor neurons. Several ALS-associated proteins, including TAR DNA-binding protein 43 (TDP-43) and Superoxide dismutase-1 (SOD-1), have been linked to small extracellular vesicles (EVs). However, the role of these EVs and their cargo in ALS patients, prior to treatment intervention, has not been investigated. This study aims to identify the earliest protein changes facilitated by EVs in ALS by examining the serum of recently diagnosed ALS patients. EVs were isolated from the serum of ALS (n = 25) and healthy control (HC, n = 9) patients before undergoing proteomics analysis. This resulted in the identification of a panel of 9 significantly up-regulated proteins and included haptoglobin and hemoglobin subunits, complement, and afamin, which are involved in pathways including heme homeostasis and autophagy. The identification of haptoglobin in ALS serum EVs suggests it has potential as an early diagnostic biomarker whilst activation of autophagy pathways suggests early recruitment of clearance pathways in ALS. This study uncovers the processes and proteins facilitated through small EVs in the initial stages of ALS. Proteomics data are available via ProteomeXchange with identifier PXD036652.

**Statement of significance of the study:** The role of small EVs, which are involved in cell-to-cell communication, and their cargo in the initiation of ALS has not been investigated. This study is the first to identify the earliest protein changes occurring in ALS through small EV facilitation. This study examined serum from newly diagnosed ALS patients, prior to treatment intervention. Therefore, the EVs, isolated from ALS and healthy control patients, captured novel ALS associated changes without confoundment from medication, which could mask early changes. A panel of 9 statistically up-regulated proteins was identified after mass spectrometry analysis. These included: haptoglobin and hemoglobin subunits, complement, and afamin. The identification of up-regulated levels of these proteins in the ALS serum EVs suggests they have potential as diagnostic biomarkers whilst identifying pathways including chaperone mediated autophagy (CMA) and microautophagy suggests early recruitment of clearance pathways in ALS. Therefore, this study uncovered the proteins being facilitated through small EVs in the initial stages of ALS.

## Introduction

Amyotrophic Lateral Sclerosis (ALS) is a fatal neurodegenerative disease characterized by the progressive accumulation of insoluble protein inclusions in upper and lower motor neurons resulting from the selective death of motor neurons in the brain and spinal cord. Motor neuron loss results in muscle atrophy, weakness and paralysis, leading to death typically within 3–5 years from ALS onset (Taylor et al., 2002). These protein aggregates accumulate in affected neurons forming Lewy body-like hyaline, skein-like inclusions, Bunina bodies, or stress granules (Mizusawa, 1993; Arai et al., 2006; Neumann et al., 2006; Miki et al., 2018; Vassileff et al., 2020). The most common proteins to form these insoluble aggregates are TAR DNA-binding protein 43 (TDP-43, encoded by *TARDBP*), a DNA and RNA binding protein involved in RNA processing, whose accumulation has been observed in 97% of sporadic and familial ALS cases, and Superoxide dismutase 1 (SOD1), an antioxidant enzyme (de Boer et al., 2021). Despite being classified as a motor neuron disease, the pathological spread of ALS begins in the primary motor cortex before progressing to the lower motor neurons and subcortical structures making it a multi-system neurodegenerative disease that begins in the brain (Brettschneider et al., 2013; Verde et al., 2017).

The complex multi-system nature of ALS creates difficulties in diagnosing the disease as no single assessment can provide a definitive early diagnosis. ALS is currently diagnosed based on the Revised Escorial and Awaji-Shima criteria which require evidence of lower and upper motor neuron degeneration in addition to signs of disease progression and spread (Brooks, 1994; Brooks et al., 2000; Carvalho and Swash, 2009). This is usually achieved through neuroimaging including Magnetic Resonance Imaging, pathological testing, nerve conduction and needle electromyography assessments (Agosta et al., 2015; Štětkářová and Ehler, 2021). Despite many assessments, ALS is commonly misdiagnosed due to overlapping symptoms with diseases including spinal-bulbar atrophy and myasthenia gravis (Jacobson et al., 2016; Singh et al., 2018). Therefore, the diagnosis of ALS heavily relies on eliminating other conditions, which often leads to a late diagnosis and treatment intervention (Štětkářová and Ehler, 2021). A definitive early blood-based biomarker would overcome this issue. Protein biomarkers such as cytokines have had varying success whilst neurofilament heavy and light chains are more effective at monitoring disease progression than diagnosis (Gendron et al., 2017; Moreno-Martinez et al., 2019; Verde et al., 2019). Recently, haptoglobin has shown promise as an early biomarker for ALS, with its early elevation correlating with disease progression and severity (Steenblock and Ikrar, 2019). Despite advancements in the biomarker field, their stability in the bloodstream is still of concern. However, this can be overcome by examining extracellular vesicle (EV) associated proteins in the blood.

Extracellular vesicles (EVs) are membranous vesicles released by all cells and encompass exosomes or small EVs, which originate from the endosomal trafficking system and are involved in inter-cellular communication (Pan et al., 1985; Kalra et al., 2012; Stotz et al., 2021). Small EVs are formed when early endosomes containing endocytosed material develop into late endosomes (Stoorvogel et al., 1991). Proteins, RNA, and lipids are subsequently transferred to the limiting membrane of the multivesicular body (MVB), causing inward budding and the generation of intra-luminal vesicles (Pan et al., 1985; Skotland et al., 2017). The MVB then either degrades the intra-luminal vesicles through fusion with the lysosomes or releases them as exosomes or small EVs upon fusion with the plasma membrane (Buschow et al., 2009). The released small EVs can then traverse long distances, including crossing the blood brain barrier, to be taken up by their target cells where they can alter the cell’s physiology (Gomes et al., 2007; Escrevente et al., 2011; Chen et al., 2016; Matsumoto et al., 2017). This unique biogenesis process enables small EVs to capture the physiological state of their parental cell, therefore, encapsulating any early disease-related changes that may be occurring.

Despite multiple protein biomarkers being investigated for diagnosing ALS, there was still no promising biomarker candidate. In this study we identify a promising panel of proteins, enriched in ALS serum EVs compared to HC serum EVs, involved in heme homeostasis and autophagy pathways, including haptoglobin. Additionally, these statistically significant proteins elucidate novel changes occurring in the early stages of ALS. This study is the first to explore the diagnostic potential of EVs before treatment intervention. Furthermore, this study presents a novel panel of biomarkers for the early diagnosis of ALS and uncovering the pathways that may be deregulated through EV facilitation in the initial stages of ALS.

## Materials and methods

### Extracellular vesicle isolation from serum

ALS and HC human serum samples were kindly supplied by Drs. Emilien Bernard, Pascal Leblanc, Anthony Fourié and Isabelle Quadrio (Human ethics: HCL.2016.710) (Lyon University Hospital, Lyon , France). The extracellular vesicles were isolated from the serum samples using the Norgen Plasma/Serum Exosome Purification Mini Kit (NOR-57400, Norgen) according to the manufacturer’s instructions. Briefly, ExoC buffer, nuclease free water, and Slurry E were added to 1 mL of serum. The sample was incubated at room temperature for 5 minutes prior to centrifugation at 2000 rpm for 2 minutes. ExoR buffer was then added to the pellet and incubated at room temperature for 5 minutes prior to centrifugation at 500 rpm for 2 minutes. The supernatant was then centrifuged through a Mini Filter Spin column at 6000 rpm for 1 minute, with the serum EVs eluting in the flowthrough.

### SDS-PAGE western blotting

The total protein content of each serum EV sample was determined using a bicinchoninic acid (BCA) protein assay (Pierce, ThermoFisher Scientific) followed by measurement using the ClarioStar microplate reader (BMG Labtech) according to manufacturer’s instructions. Samples were lysed (5M NaCl, 1M Tris, Triton X-100, 1% (w/v) sodium deoxycholate, 1x cOmplete ULTRA protease inhibitor) and incubated at 4°C for 20 minutes prior to centrifugation at 2,500 x g at room temperature for 5 minutes. The supernatant was combined with 6X sodium dodecyl sulphate (SDS) loading dye (0.375 M Tris, 12% SDS, 30% glycerol, 0.06% Bromophenol blue, pH 6.8), containing 5% β-mercaptoethanol, and incubated at 95°C for 5 minutes. Electrophoresis was performed on a 4-12% Bis-Tris Plus Gel (NuPAGE or Bolt, Invitrogen) with 1X MES SDS running buffer (NuPAGE, Invitrogen). The proteins were then transferred to PVDF membrane and probed with the following antibodies (Actin, Cell Signalling 8H10D10; Flotillin-1, BD Bioscience 610821; Tsg101, Abcam ab83; Nucleoporin, BD Bioscience 610497; Apolipoprotein B (EPR2914), Abcam ab139401) in 2.5% skim milk in PBS-T (0.05% Tween). The membranes were then washed in PBS-T (0.05% Tween) and probed with either mouse IgG HRP (BioStrategy NA931) or rabbit IgG HRP (BioStrategy NA934) secondary antibody before development with Clarity ECL reagent (Bio-Rad) and imaged using the ChemiDoc Touch imaging system (Bio-Rad) as per the manufacturer’s recommendations. Image Lab 5.2.1 (Bio-Rad) was used to analyze the images.

### Nanoparticle tracking analysis

Isolated vesicles underwent Nanoparticle Tracking Analysis to determine their size and concentration. The samples were diluted 1 in 1000 in filtered and degassed Dulbecco’s phosphate-buffered saline before being infused through a 1 mL syringe into the ZetaView© Quatt PMX-420 (Particle Metrix). Recordings were created by scanning eleven positions within the instrument’s cell, each capturing 30 frames per position using the following parameters: Maximum particle size: 1000, Minimum particle size 10, Minimum Brightness 25, Focus: autofocus, Sensitivity: 80.0, Shutter: 100, and Cell temperature: 25°C. These recordings were analyzed using the in-built ZetaView Software 8.05.14-SP7 to determine the size and concentration of the vesicles.

### Transmission electron microscopy

Isolated vesicles were observed using Transmission Electron Microscopy to determine their size and morphology. Following glow discharge for 60 seconds, a formvar-copper coated grid (ProSciTech) was loaded with 5 μL of sample and incubated at room temperature for 30 seconds. The excess sample was blotted off and 5 μL of Uranyl acetate (Agar Scientific) was applied to the grid for 10 seconds, twice. The grid was then imaged using the JEM-2100 Transmission Electron Microscope (Jeol).

### Protein digest and peptide desalting

To denature protein, samples were reconstituted in urea (8 M urea, 25 mM Tris-HCl, pH 8.0). Disulphide bonds were reduced by the addition of TCEP (tris-2-carboxyethyl-phosphine) to 2 mM (60 minute incubation) followed by the addition of iodoacetamide to 38 mM (45 minute incubation, in the dark) to alkylate reduced thiols. The samples were then diluted with 20 mM Tris-HCl to reduce urea concentration below 1 M before the addition of sequencing grade trypsin (Promega) to achieve a 1:50 ratio compared to the original protein amount. Trypsin digestion was completed overnight at 37 °C. Tryptic peptides were desalted and concentrated using StageTips according to the published protocol (Rappsilber et al., 2007).

### LC-MS analysis of peptides

LC-MS was performed on a Thermo Ultimate 3000 RSLCnano UHPLC system and a Thermo Orbitrap Eclipse Tribrid mass spectrometer. Peptides were reconstituted in 0.1% (v/v) trifluoroacetic acid (TFA), 2% (v/v) acetonitrile (ACN), and 500 ng of peptides were loaded onto PepMap C18 5 µm 1 cm trapping cartridge (Thermo-Fisher Scientific, Waltham, MA, USA) at 12 µL/min for 6 minutes and washed for 6 minutes before switching the pre-column in line with the analytical column (nanoEase M/Z Peptide BEH C18 Column, 1.7 µm, 130 Å and 75 µm ID × 25 cm, Waters). The separation of peptides was performed at 250 nL/min using a linear ACN gradient of buffer A (0.1% (v/v) formic acid, 2% (v/v) ACN) and buffer B (0.1% (v/v) formic acid, 80% (v/v) ACN), starting at 14% buffer B to 35% over 90 minutes, then rising to 50% B over 15 minutes followed by 95% B in 5 minutes. The column was then cleaned for 5 minutes at 95% B and afterwards a 3 minute equilibration step at 1% B. Data were collected in Data Dependent Acquisition mode using m/z 350–1500 as MS scan range and HCD MS/MS spectra were collected by the orbitrap using a cycle time of 3 seconds per MS scan at 30,000 resolution. Dynamic exclusion parameters were set as follows: exclude isotope on, duration 60 seconds and using the peptide monoisotopic peak determination mode. Other instrument parameters for the instrument were: MS scan at 120,000 resolution, injection time Auto, AGC target Standard, HCD collision energy 30%, injection time Auto with AGT target at Standard. The isolation window of the quadrupole for the precursor was 1.6 m/z. Easy-IC internal mass calibration was used.

### Proteomics database search

LC-MS raw files were searched using Thermo Proteome Discoverer 2.4 and the sequest HT search versus the Uniprot Homo sapiens reference proteome (29 Jan 2021) with reversed decoy sequences. Precursor tolerance was set to 10 ppm and fragment tolerance to 0.05 Da. Three missed trypsin cleavages were permitted and max peptide length of 144 aa. The included static modification was carbamidomethyl of C, and dynamic modifications were oxidation of M, acetylation and/or methionine loss of protein N-terminus. Validation FDR thresholds were 0.01 for strict or 0.05 for relaxed criteria at PSM, peptide and protein levels. Label-free quantification was performed on precursor ion intensity and abundances were normalized to total peptide amount. Abundance values were analyzed using Perseus (1.6.15.0). Abundances were log(2) transformed before imputation using Perseus replace missing values from a normal distribution function (width 0.2, downshift 1.9, per column). Students T-test was used to compare ALS vs HC.

The mass spectrometry proteomics data have been deposited to the ProteomeXchange Consortium via the PRIDE (Perez-Riverol et al., 2022) partner repository with the dataset identifier PXD036652.

Access details for reviewers. **Username:**. **Password:** FgTI6SPx

### Bioinformatics

The statistically significantly differentially expressed proteins were then analyzed using Funrich3.1.3 to determine the molecular function, biological processes, and cellular component distribution of the panel of biomarkers (Pathan et al., 2015). The proteins also underwent reactome analysis to determine the pathway involvement of the biomarkers (Gillespie et al., 2022). Finally, STRING analysis was utilized to determine whether any of the biomarkers are interacting partners (Szklarczyk et al., 2021).

## Results

### Serum EVs were characterized as small EVs

The Norgen Plasma/Serum Exosome Purification Mini Kit was characterized using human serum samples from the Red Cross. The isolated vesicles underwent western blot analysis where they were probed for EV enriched and non-EV enriched markers to ensure the presence of small EVs and the absence of contamination (**Figure 1A**). The size of the vesicles was determined to be consistent with that of small EVs through NTA analysis, performed on the ZetaView© Quatt PMX-420 (**Figure 1B**) (Sokolova et al., 2011; Kalra et al., 2012). Furthermore, the vesicles were shown to consist of a homogenous population exhibiting characteristic EV features including 40-120 nm diameters (**Figure 1C**) (Wu et al., 2015). Following successful characterization of the kit, EVs were isolated from human ALS (n = 25) and HC (n = 9) serum samples, the sample demographics of which are detailed in **Table 1**. Following successful isolation using the Norgen Plasma/Serum Exosome Purification Mini Kit, the protein content of serum EVs was determined using LC-MS proteomic analysis.

**Figure 1.**
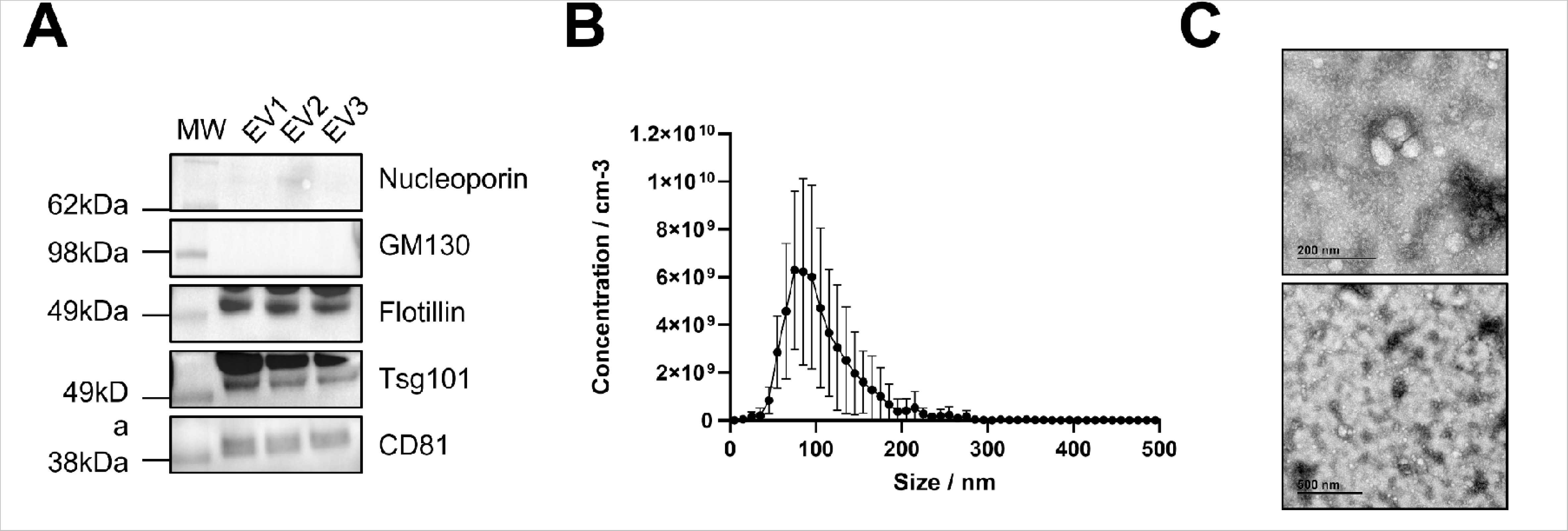
Characterisation serum extracellular vesicles (EVs). Vesicles isolated from the red cross serum samples demonstrate the characteristics of small extracellular vesicles (EVs). **(A).** The serum EVs expressed small EV enriched markers tsg101, flotillin, and CD81, and the absence of small EV non-enriched markers; GM130 and nucleoporin. The blot is representative of n = 30. **(B).** Nanoparticle Tracking Analysis of the serum EVs analysed by the ZetaView© Quatt PMX-420 showed the appearance of vesicles around 80 nm in diameter, a size consistent with that of small EVs such as exosomes. This graph is of a representative of n = 3. **(C).** Transmission Electron Microscopy images of the serum EVs display a population of small EVs 40–100 nm in diameter, with depressed cup-like structures and double lipid membranes. The images are from a representative subject.

**Table I:** Demographic of human Amyotrophic Lateral Sclerosis (ALS) and control serum samples.

| Sample Information |  |  |  |  |  |  |
| --- | --- | --- | --- | --- | --- | --- |
| ID | Sex | Diagnostic | Age | Mutation status | site of onset | FTD status |
| 1 | F | ALS | 66-75 | *C9ORF72* positive | lower limb | Yes |
| 2 | F | ALS | 56-65 | *C9ORF72* positive | bulbar | Yes |
| 3 | M | ALS | 56-65 | *C9ORF72* negative | lower limb | N/A |
| 4 | F | ALS | 76-85 | Pathological mutation<br>*TARDBP* | upper limb | N/A |
| 5 | M | ALS | 66-75 | *C9ORF72* negative | bulbar | N/A |
| 6 | M | ALS | 56-65 | *C9ORF72* negative | lower limb | N/A |
| 7 | F | ALS | 66-75 | *C9ORF72* negative | bulbar | N/A |
| 8 | F | ALS | 56-65 | *C9ORF72* positive | upper limb | Yes |
| 9 | M | ALS | 56-65 | *C9ORF72* negative | lower limb | N/A |
| 10 | M | ALS | 76-85 | *C9ORF72* negative | lower limb | N/A |
| 11 | M | ALS | 56-65 | *C9ORF72* negative | lower limb | N/A |
| 12 | M | ALS | 66-75 | *C9ORF72* negative | lower limb | N/A |
| 14 | F | ALS | 56-65 | Pathological mutation<br>*TARDBP* | upper limb | N/A |
| 15 | M | ALS | 56-65 | *C9ORF72* positive | lower limb | Yes |
| 16 | F | ALS | 66-75 | *C9ORF72* negative | upper limb | Yes |
| 17 | M | ALS | 66-75 | *C9ORF72* negative | bulbar | N/A |
| 18 | M | ALS | 56-65 | *C9ORF72* negative | lower limb | N/A |
| 19 | M | ALS | 56-65 | *C9ORF72* negative | upper limb | N/A |
| 20 | F | ALS | 66-75 | *C9ORF72* negative | bulbar | Yes |
| 21 | F | ALS | 76-85 | Pathological mutation<br>*TARDBP* | upper limb | N/A |
| 22 | M | ALS | 66-75 | *C9ORF72* negative | bulbar | N/A |
| 23 | M | ALS | 46-55 | Unknown | upper limb | N/A |
| 24 | M | ALS | 85+ | *C9ORF72* negative | bulbar | Yes |
| 25 | M | ALS | 76-85 | *C9ORF72* negative | bulbar | N/A |
| 28 | M | Control | 46-55 | N/A | N/A | N/A |
| 29 | F | Control | 56-65 | N/A | N/A | N/A |
| 30 | F | Control | 25-35 | N/A | N/A | N/A |
| 31 | M | Control | 25-35 | N/A | N/A | N/A |
| 32 | M | Control | 25-35 | N/A | N/A | N/A |
| 33 | F | Control | 56-65 | N/A | N/A | N/A |
| 34 | F | Control | 36-45 | N/A | N/A | N/A |
| 35 | M | Control | 36-45 | N/A | N/A | N/A |
| 36 | M | Control | 25-35 | N/A | N/A | N/A |
| 37 | M | ALS | 56-65 | *C9ORF72* positive | lower limb | Yes |

\*FTD = Frontal Temporal Dementia

### Protein upregulation was observed in ALS serum EVs compared to HC serum EVs

The proteome of the ALS serum EVs was compared to that of the HC serum EVs, through mass spectrometry analysis, to determine whether specific proteins were differentially packaged in the disease state. There was an absence of all-encompassing or universal changes in the proteome when comparing the ALS and HC serum EVs (**Figure 2A**). The ALS samples did appear to form two groups in hierarchical clustering analysis, one group clustering with HC samples and the other composed of only ALS samples. Despite this clustering, neither group appeared to be exclusively defined by the age, diagnosis, sex, gene status, or site of onset. This could be attributed to the samples being collected from symptomatic patients before treatment initiation. However, the ALS serum EVs did appear to contain an upregulation of certain groups of proteins compared to the HC serum EVs (**Figure 2B**), suggesting a potential change in EV cargo loading associated with the early stages of ALS. Immunoglobin contaminants and inter-alpha-trypsin inhibitor were removed from further downstream analysis.

**Figure 2.**
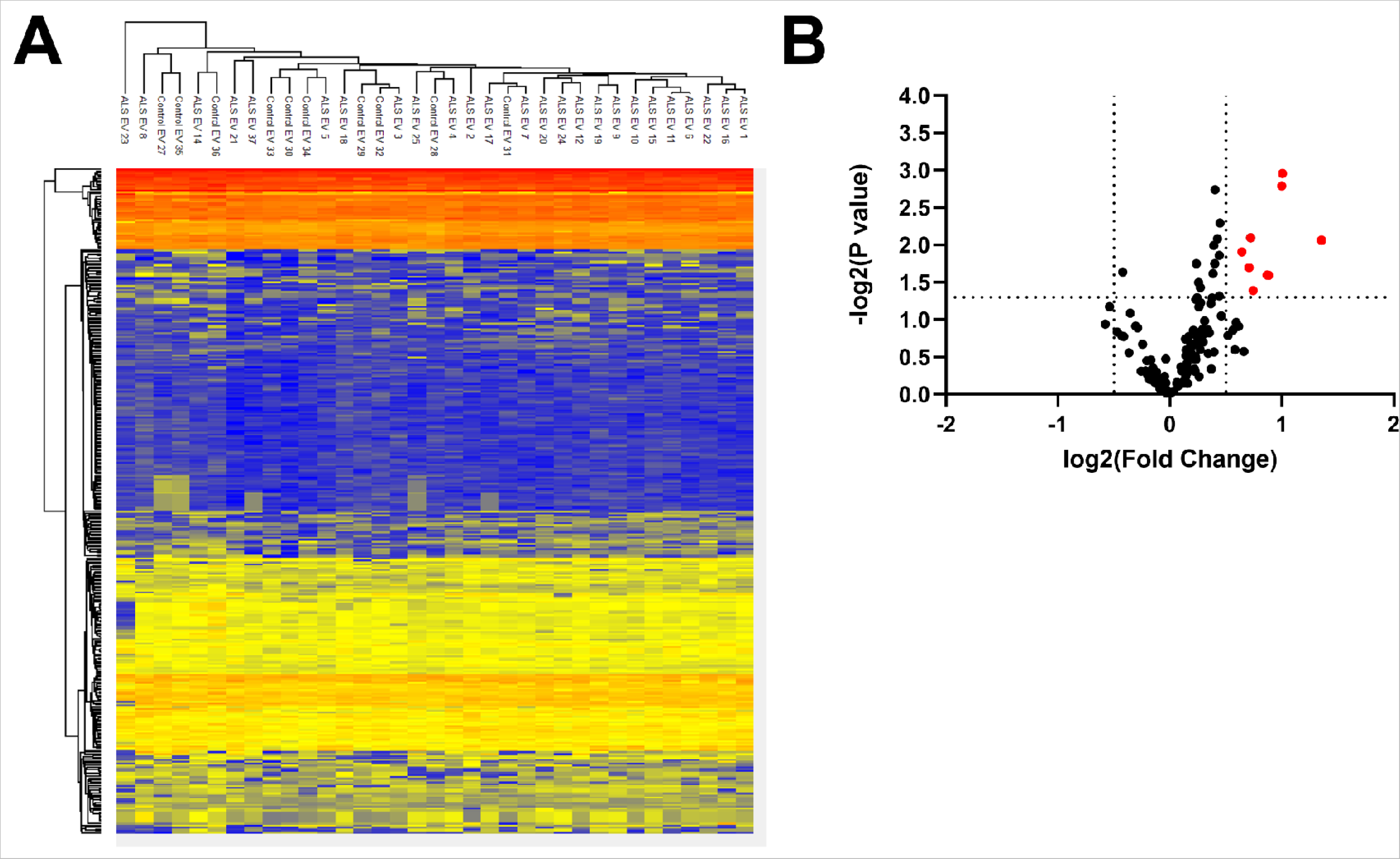
Proteome analysis of Amyotrophic Lateral Sclerosis (ALS) and healthy control (HC) serum extracellular vesicles (EVs). ALS serum EVs appear to be upregulated in protein cargo. **(A).** Heat map depicting relative abundancy of ALS serum EVs and healthy control (HC) serum EVs, which appear to have similar proteome profiles. **(B).** Volcano plot revealing the ALS serum EVs appear to be upregulated in certain groups of proteins compared to the HC serum EVs.

### Upregulated proteins in ALS serum EVs are involved in heme homeostasis and autophagy pathways

The proteins found to be statistically significantly enriched in ALS serum EVs primarily consisted of hemoglobin subunits (**Figure 3** and **Table 2**). Proteins did lose significance following FDR-correction (Table 2). The 9 enriched proteins in ALS serum EVs (**Figure 3**) underwent testing for enrichment of gene ontology (GO) terms to uncover their potential role in ALS. GO analysis revealed the differentially expressed proteins exhibiting molecular functions including transporter and complement activity. Regarding biological processes, the proteins were involved in transport and immune response. Additionally, the proteins were primarily found to be in the extracellular, the hemoglobin complex, and the lysosome cellular compartments (**Figure 4A and Supplementary Table 2**). Reactome analysis was subsequently performed on the proteins to determine which pathways their deregulation was affecting. The analysis identified the 9 statistically significantly enriched proteins in ALS serum EVs to be involved in heme signalling and scavenging, complement pathway, cellular response to chemical stress, small molecule and vesicle-mediated transportation, and chaperone-mediated autophagy and microautophagy (**Table 3** and **Figure 4B**). Given that the reactome results revealed the presence of two major pathway groups: heme homeostasis and autophagy, STRING analysis was performed to determine whether the proteins were interacting. Interestingly, all 9 statistically significantly enriched ALS serum EV proteins were determined to have some form of interaction with at least one other protein, with 8 of these proteins exhibiting well characterized interactions (**Figure 5**). Together, the GO, reactome, and STRING analysis elucidated several functions and pathways whose dysregulation may be mediated through small EVs in ALS.

**Figure 3.**
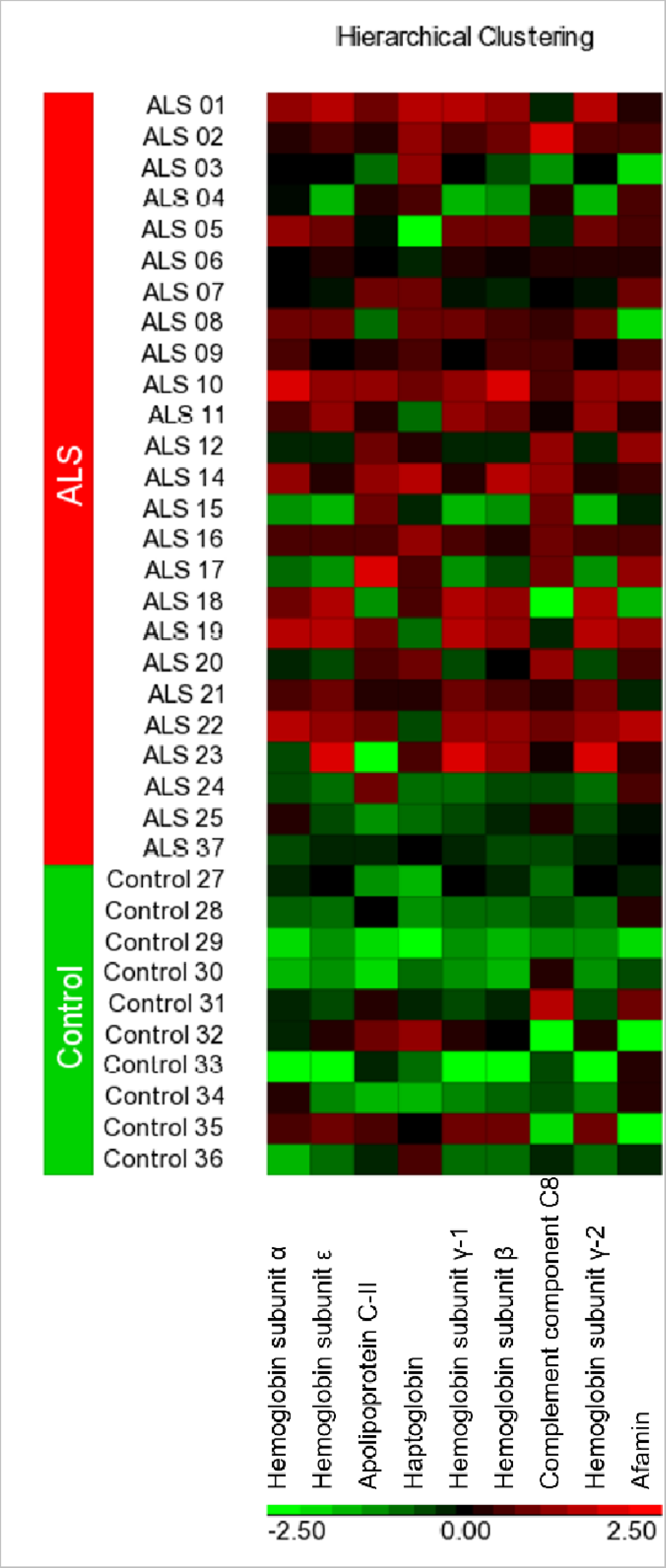
Statistically significantly differentially expressed proteins in Amyotrophic Lateral Sclerosis (ALS) vs healthy control (HC) serum extracellular vesicles (EVs). The most statistically significantly differentially expressed proteins in ALS serum EVs appear to be upregulated when compared to the HC serum EVs.

**Figure 4.**
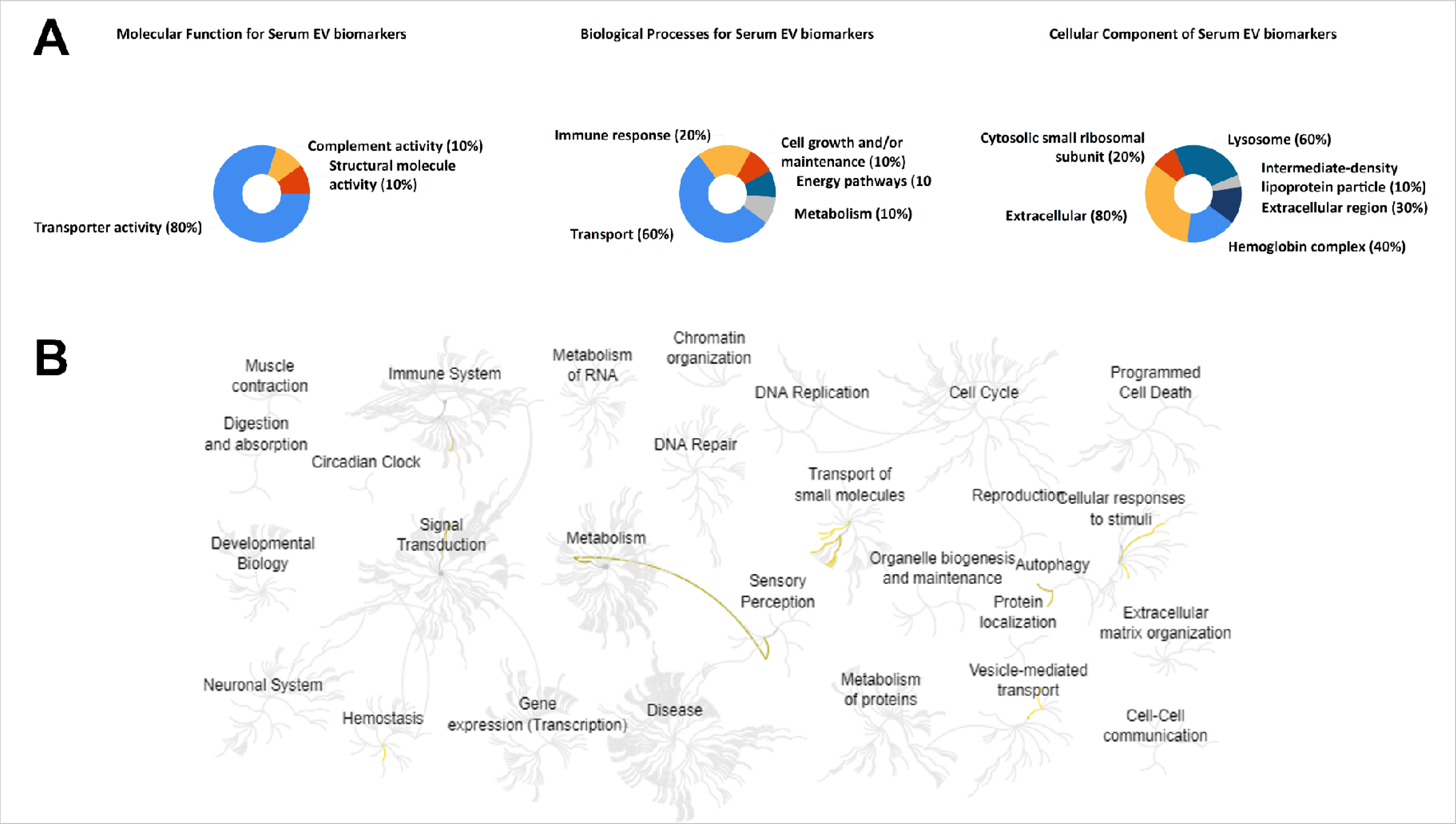
Gene ontology (GO) and reactome analysis of the 9 statistically significantly upregulated proteins. The Amyotrophic Lateral Sclerosis (ALS) serum EV upregulated proteins are involved in autophagy, transport, and heme homeostasis. **(A).** GO analysis revealed the differentially expressed proteins exhibit molecular functions including transporter activity and complement activity; biological processes including transport and immune response; and cellular component locations including the extracellular, the hemoglobin complex, and the lysosome. **(B).** Reactome analysis uncovered the upregulated proteins to be in involved in metabolism, signal transduction, immune response, transport of small molecules, cellular response to stimuli, autophagy, protein localization, and vesicle-mediated transport.

**Figure 5.**
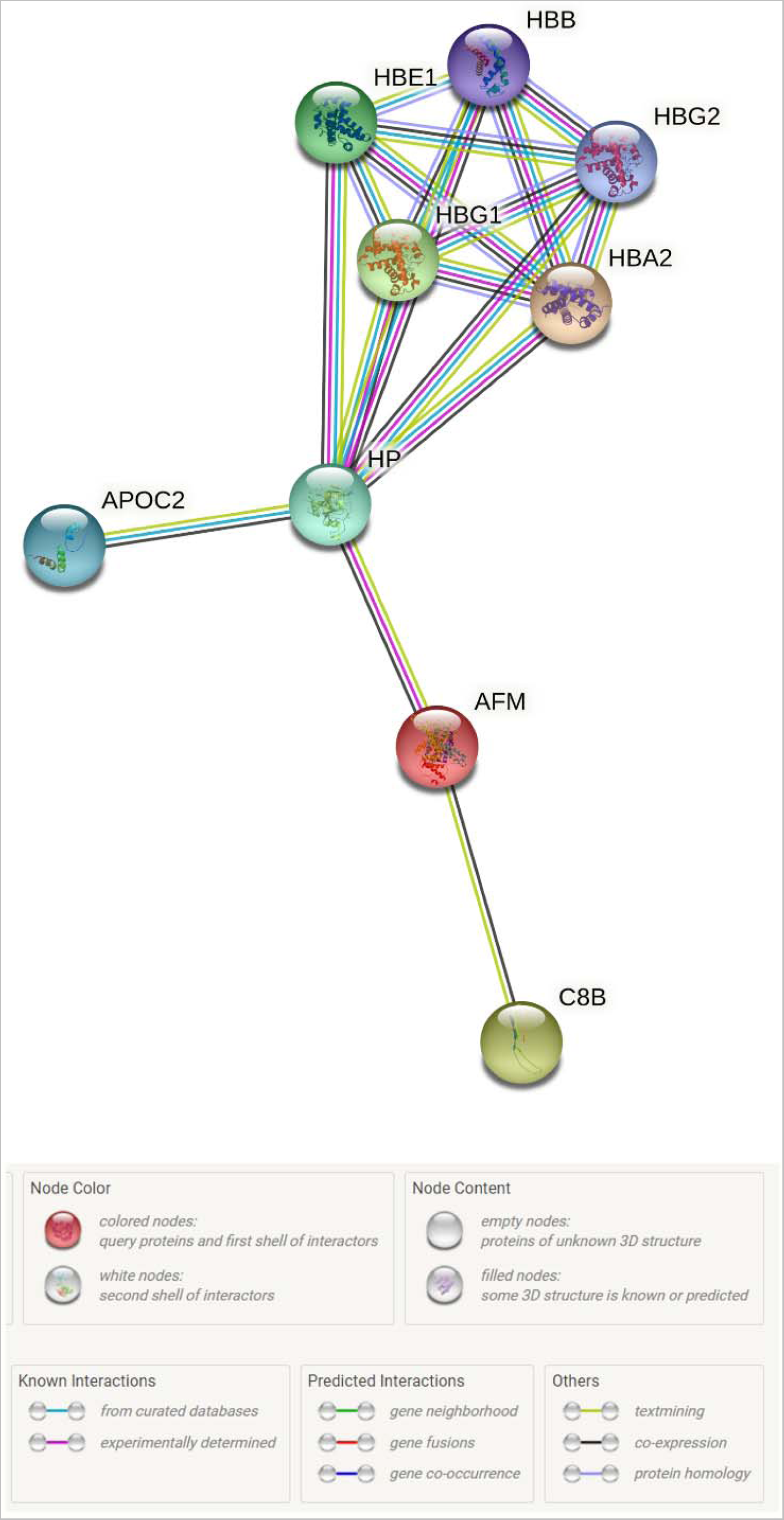
Search tool for the retrieval of interacting genes/proteins (STRING) analysis of the 9 statistically significantly upregulated proteins. All 9 statistically significantly upregulated Amyotrophic Lateral Sclerosis (ALS) serum EV proteins were determined to exhibit network relationships/connectivity or some form of interaction with at least one other protein. Graph was created using STRING.

**Table II:** Statistically significantly enriched proteins in the Amyotrophic Lateral Sclerosis (ALS) serum EVs vs. the healthy control (HC) serum extracellular vesicles (EVs)

| **Accession** | **Description** | **-Log Student's**<br>**T-test p-value**<br>**ALS_NC** | **Student's T-**<br>**test q-value**<br>**ALS_NC** | **Student's T-**<br>**test Difference**<br>**ALS_NC** |
| --- | --- | --- | --- | --- |
| P69905 | Hemoglobin subunit alpha | 2.959 | 0.268 | 1.006 |
| P68871 | Hemoglobin subunit beta | 2.791 | 0.178 | 0.998 |
| P43652 | Afamin | 1.596 | 0.173 | 0.883 |
| P02655 | Apolipoprotein C-II | 1.599 | 0.179 | 0.868 |
| P02100 | Hemoglobin subunit epsilon | 2.097 | 0.282 | 0.721 |
| P69891 | Hemoglobin subunit gamma-1 | 2.097 | 0.235 | 0.721 |
| P69892 | Hemoglobin subunit gamma-2 | 2.097 | 0.202 | 0.721 |
| P07358 | Complement component C8 beta chain | 1.697 | 0.168 | 0.708 |
| P00738 | Haptoglobin | 1.909 | 0.134 | 0.643 |

**Table III:**
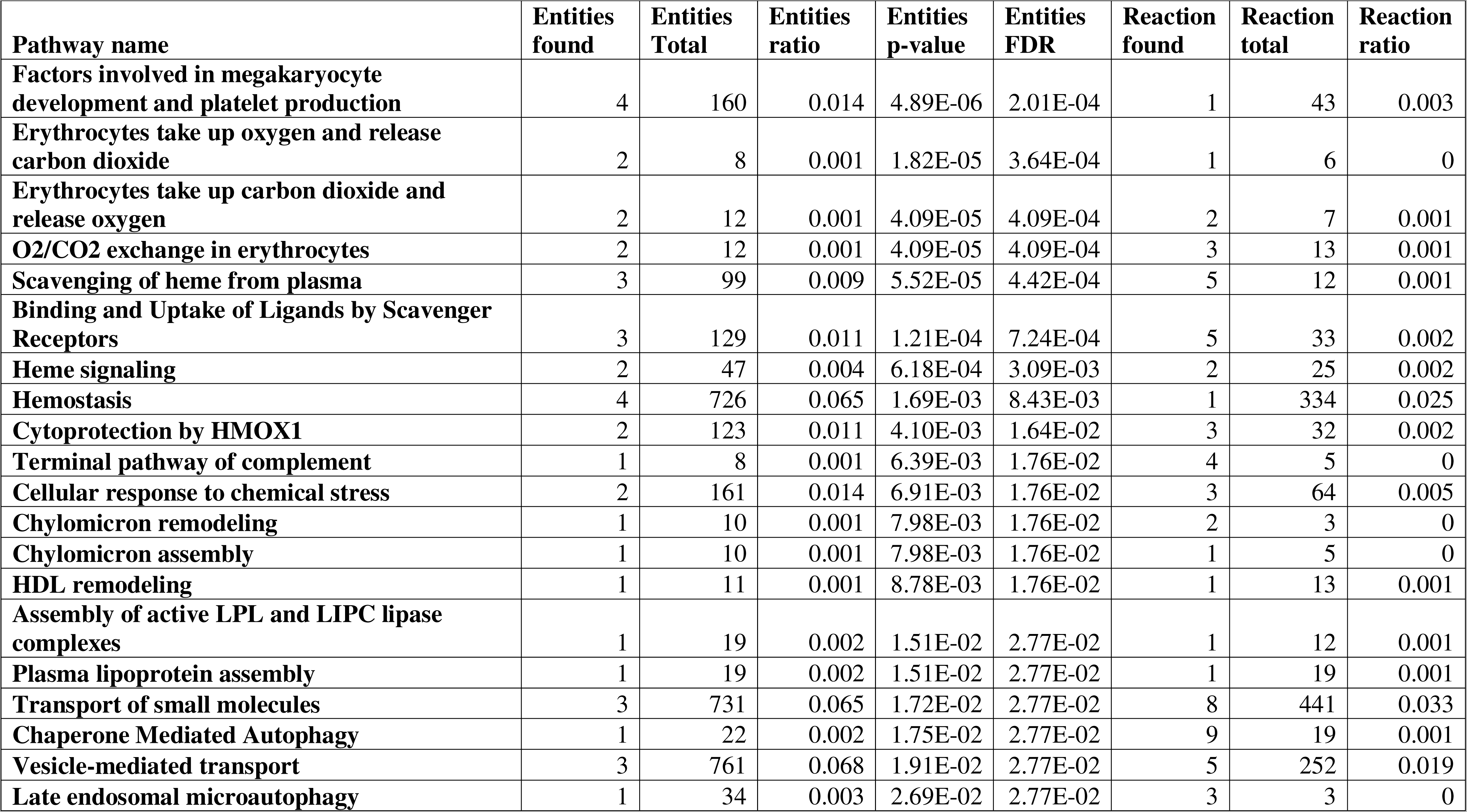
Pathways affected by the statistically significantly enriched proteins in the Amyotrophic Lateral Sclerosis (ALS) serum extracellular vesicles (EVs)

## Discussion

This study identified 9 statistically significantly upregulated proteins in ALS serum EVs compared to the HC serum EVs, potentially revealing a group of biomarkers for a liquid biopsy for ALS. These proteins were found to function in heme homeostasis and autophagy pathways revealing pathways potentially deregulated in ALS. Most importantly, these samples were collected from symptomatic patients before any treatment administration. Therefore, the ALS serum EVs isolated and examined in this study have potentially captured changes occurring in ALS without confoundment from treatments.

An early detection test for diagnosing ALS is vital, with treatments, including Riluzole, being proven most effective when administered early and for prolonged periods (Zoccolella et al., 2007; Fang et al., 2018). Therefore, a blood test targeting early protein changes, protected in EVs from enzymes in the bloodstream, could address this issue. The serum EVs were isolated using a commercial kit which is known to co-isolate lipoproteins hence accounting for the lipoprotein positive marker detected in the EV samples. Despite the presence of lipoprotein, the samples are EV-enriched as indicated by the presence small EV markers, thus the samples are referred as serum EVs. The ALS serum EVs isolated from newly diagnosed symptomatic patients contained 9 statistically significantly upregulated proteins, which could serve as biomarkers for the diagnosis of ALS. Of these proteins, haptoglobin was detected. Haptoglobin is involved in binding free hemoglobin released from erythrocytes. This binding inhibits the iron in hemoglobin from undergoing autoxidation and producing superoxide which is processed by SOD1 to produce hydrogen peroxide, an oxidative agent (Rifkind et al., 2015). Once bound, this extracellular complex is abruptly cleared to prevent an increase in free circulating iron (Rifkind et al., 2015). Increased iron has been detected in ALS patients’ motor cortex, cerebral spinal fluid, and serum (Ignjatović et al., 2013; Veyrat-Durebex et al., 2014; Zheng et al., 2017; Sanghai and Tranmer, 2021). Additionally, haptoglobin has not only been detected in EVs but, has been found to be elevated in EVs isolated from the plasma of cancer patients and supernatant of cell culture cancer models (Zheng et al., 2020; Priya et al., 2022). The detection of increased EV haptoglobin in an in vitro setting suggests haptoglobin is not a contaminant and its packaging into EVs is being disrupted in disease settings (Zheng et al., 2020; Priya et al., 2022). The detection of elevated levels of hemoglobin subunits and the identification of the hemoglobin complex in the GO’s cellular component analysis of the EVs, suggests an increased extracellular iron concentration consistent with the literature (Yu et al., 2018). Increased hemoglobin and heme in the brain, as observed in ALS, is known to promote oxidative stress, inflammation, lipid peroxidation, and mitochondrial dysfunction which has been linked to initiating neurodegeneration (Shi et al., 2010; Righy et al., 2016; Chiabrando et al., 2018). This suggests the increase in haptoglobin detected in ALS serum EVs may be an early attempt to prevent further free radical production and neurodegeneration. However, given these EVs were isolated from blood samples, the possibility of hemolysis cannot be eliminated, especially given the samples were shipped from overseas. In order to, verify whether the elevated hemoglobin and haptoglobin levels are due to hemolysis this study will need to be repeated in an independent cohort with new samples. Haptoglobin has been suggested to play a role in immune regulation (Steenblock and Ikrar, 2019). Given immune response was a biological process identified in the GO analysis, our study further supports haptoglobin’s immunomodulatory role in the early stages of ALS. Finally, serum haptoglobin levels were discovered to positively correlate with sporadic ALS, disease stage, and be predictive of progression rate (Steenblock and Ikrar, 2019). Therefore, the upregulation of haptoglobin in ALS serum EVs elucidates dysregulation that may be occurring in the disease prior to treatment.

The detection of complement component C8 beta chain, which is involved in forming the membrane attack complex (MAC), may reveal another pathway dysregulated in ALS. Furthermore, complement activity was determined to be a molecular function based on the GO’s cellular function analysis. During an immune response MAC inserts into the target cell’s membrane to form a pore through which water and ions enter leading to osmotic lysis (Morgan, 2016). MAC was found on neuromuscular junctions of intercostal muscles from ALS patients, in addition to the spinal cord and motor cortex of ALS patients (Sta et al., 2011; Bahia El Idrissi et al., 2016). This late complement component activation was determined to be significantly higher on microglia in ALS tissues compared to controls (Sta et al., 2011). Additionally, elevated levels of MAC components including C8 have been detected in ALS patient plasma (Mantovani et al., 2014). Together these results suggest that terminal complement activation may be occurring in the early stages of ALS before denervation and may be one of the initial pathways activated in ALS pathogenesis. Another protein found to be statistically significantly upregulated in ALS serum EVs was Afamin, a vitamin E binding glycoprotein. Interestingly, vitamin E supplementation has been linked to lower incidences and lower risk of dying from ALS (Ascherio et al., 2005; Veldink et al., 2007; Wang et al., 2011; D’Amico et al., 2021). These results suggest an increase in Afamin could indicate an increase in vitamin E to counteract the dysfunction occurring during the early stages of ALS. However, a recent study discovered higher vitamin E levels in ALS patients and a positive correlation between vitamin E and an increased risk for ALS (Wang et al., 2020). Therefore, the upregulation of Afamin in ALS serum EVs is complicated by the confounding literature regarding vitamin E in ALS.

Contrastingly, dysfunction of autophagy pathways is well documented in ALS. Both chaperone mediated autophagy and late endosomal microautophagy were determined to be affected through reactome analysis as a result of hemoglobin subunit beta. Chaperone mediated autophagy (CMA) is involved in the removal of damaged and aggregated proteins (Boland et al., 2018; Arosio et al., 2020). CMA begins when the Hsc70 chaperone recognizes the soluble protein’s specific KFERQ pentapeptide, to which the chaperone binds and translocates the complex to the lysosome (Wing et al., 1991; Kaushik and Cuervo, 2018). Upon reaching the lysosome, the protein-Hsc70 complex binds to the lysosomal associated membrane protein 2A (Lamp2A), which forms a pore through which the protein may enter the lysosome for degradation (Kaushik and Cuervo, 2018). Aggregated forms of TDP-43 have been found to bind Hsc70 and co-localize with Lamp2A in CMA lysosomes where they elicit lysosomal damage (Ormeño et al., 2020). Additionally, reduced Hsc70 expression and increased insoluble TDP-43 levels were found in ALS patients peripheral blood mononuclear cells (Arosio et al., 2020). Therefore, the discovery of CMA in the reactome analysis suggests the cells may be upregulating this pathway to remove any newly aggregated or damaged proteins including TDP-43, in the early stages of ALS. Alternatively, microautophagy involves the internalization of cytosolic proteins and organelles by late endosomal and lysosomal vesicles (Vicencio et al., 2020). However, like CMA, microautophagy can selectively target proteins with the KFERQ pentapeptides through recruitment of HSPA8 and Hsc70 chaperones (Li et al., 2012; Tekirdag and Cuervo, 2018). Additionally, the lysosome constituted a large proportion of the GO’s cellular component analysis, further alluding to the activation of clearance pathways. Identifying vesicle-mediated transport in the reactome analysis suggests the shuttling of proteins through alternative routes, including small EV packaging, upon congestion of the clearance pathways. Stress granule and RNA binding proteins were found to be packaged into small EVs upon congestion of the macroautophagy pathway in the post-mortem brain tissue of late-stage ALS subjects (Vassileff et al., 2020). Therefore, the identification of both the CMA and microautophagy pathways suggests KFERQ motif containing proteins are being packaged into the ALS EVs upon impediment of autophagy pathways in ALS.

This study was the first to examine serum EVs isolated from ALS patients prior to treatment intervention, revealing the initial changes occurring in symptomatic patients. Despite a small sample size, 9 statistically significant differentially expressed proteins were identified in ALS serum EVs compared to the control serum EVs. The proteins consisted of haptoglobin and hemoglobin subunits, complement and afamin, and activated pathways including heme homeostasis and autophagy. The identification of haptoglobin in ALS serum EVs is especially promising for the diagnosis of ALS as it has shown promise as an early biomarker for ALS in the literature. In the future, a larger sample size should enable validation of these pathways and identification of other pathways involved in the disease. Furthermore, a sample cohort consisting of ALS patients and controls closer in age would be required for the validation cohort to ensure the changes observed are not simply an artifact of age. Despite these limitations, the identification of haptoglobin in ALS serum EVs further supports its role as an early biomarker for the diagnosis of ALS. Given an absence of treatment confoundment in these samples, the upregulation of this panel of proteins makes them a very promising target in a liquid biopsy for the diagnosis of ALS. Additionally, the proteins’ involvement in CMA and microautophagy pathways suggests the proteins may be packaged into EVs upon impairment of the autophagy pathways. Together these results unveil the pathogenic changes, being facilitated through small EVs, in ALS prior to treatment initiation. This not only better elucidates disease dysregulation, it additionally provides a panel of biomarkers that can be used in the diagnosis of suspected ALS patients.

## Supplementary Materials

**Supplementary Table S1:** LFQ values, p-values, fold change, and relative abundance values of all proteins detected in the Amyotrophic Lateral Sclerosis (ALS) and healthy control (HC) serum EVs.

**Supplementary Table S2:**
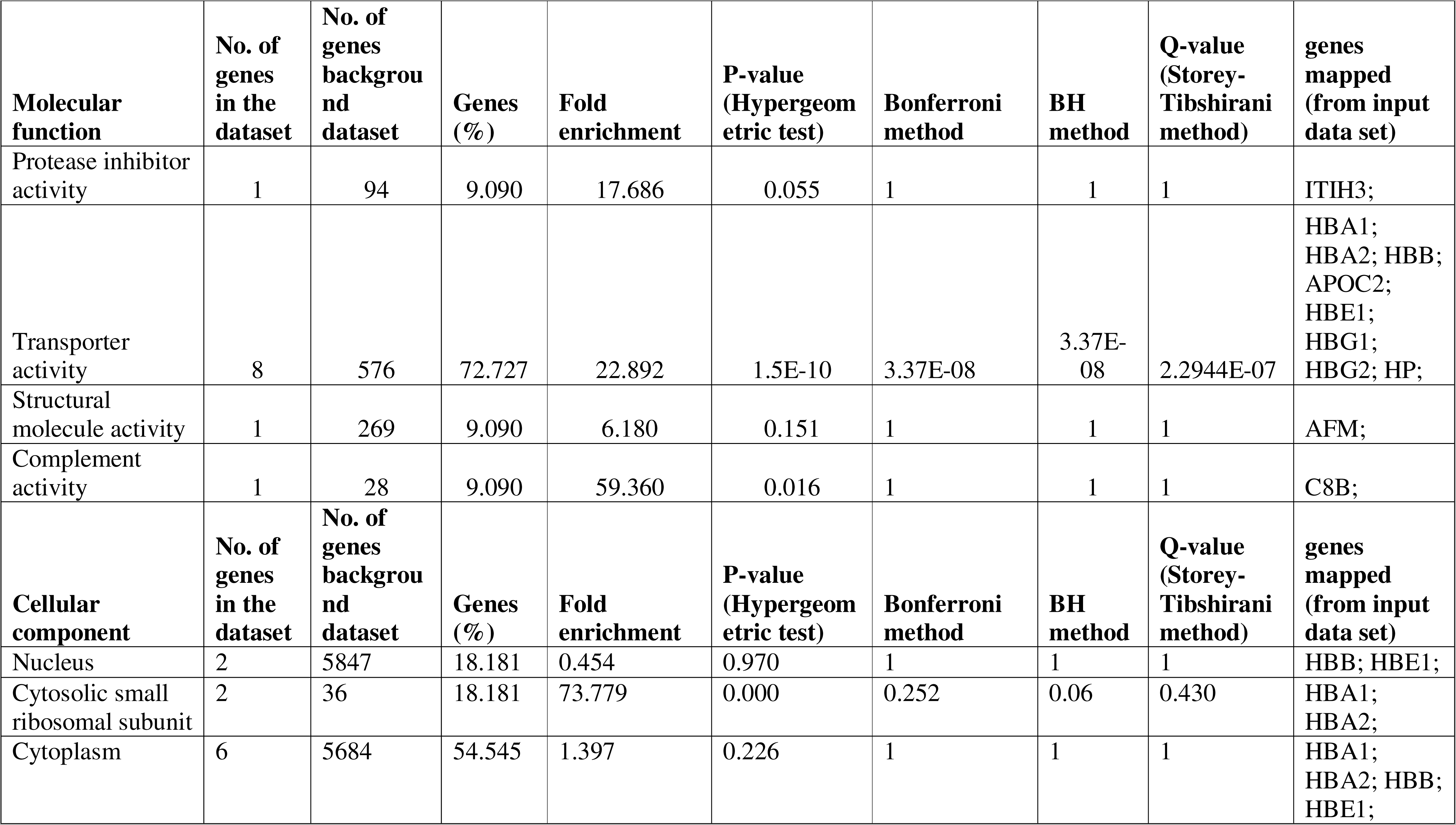
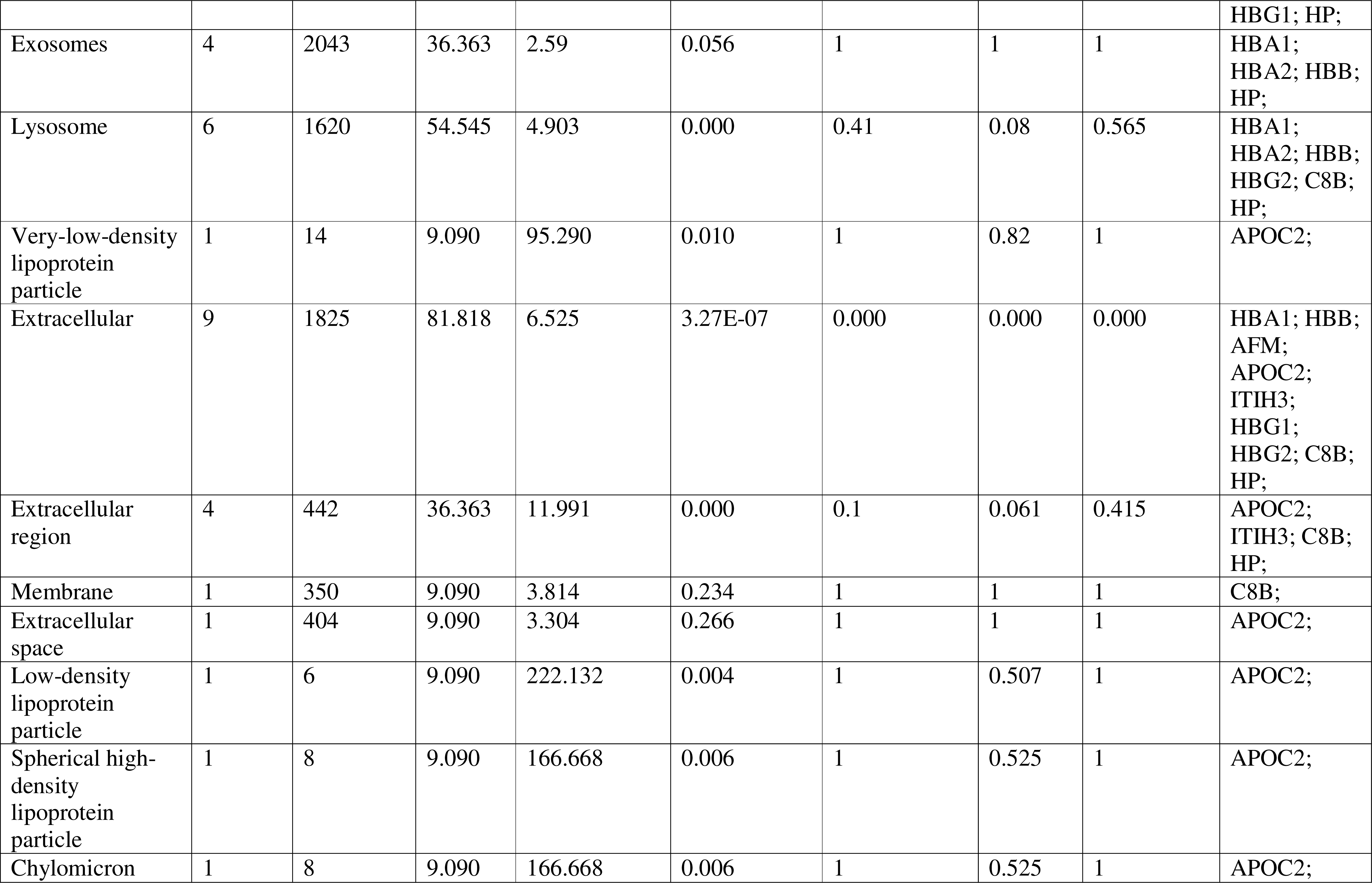
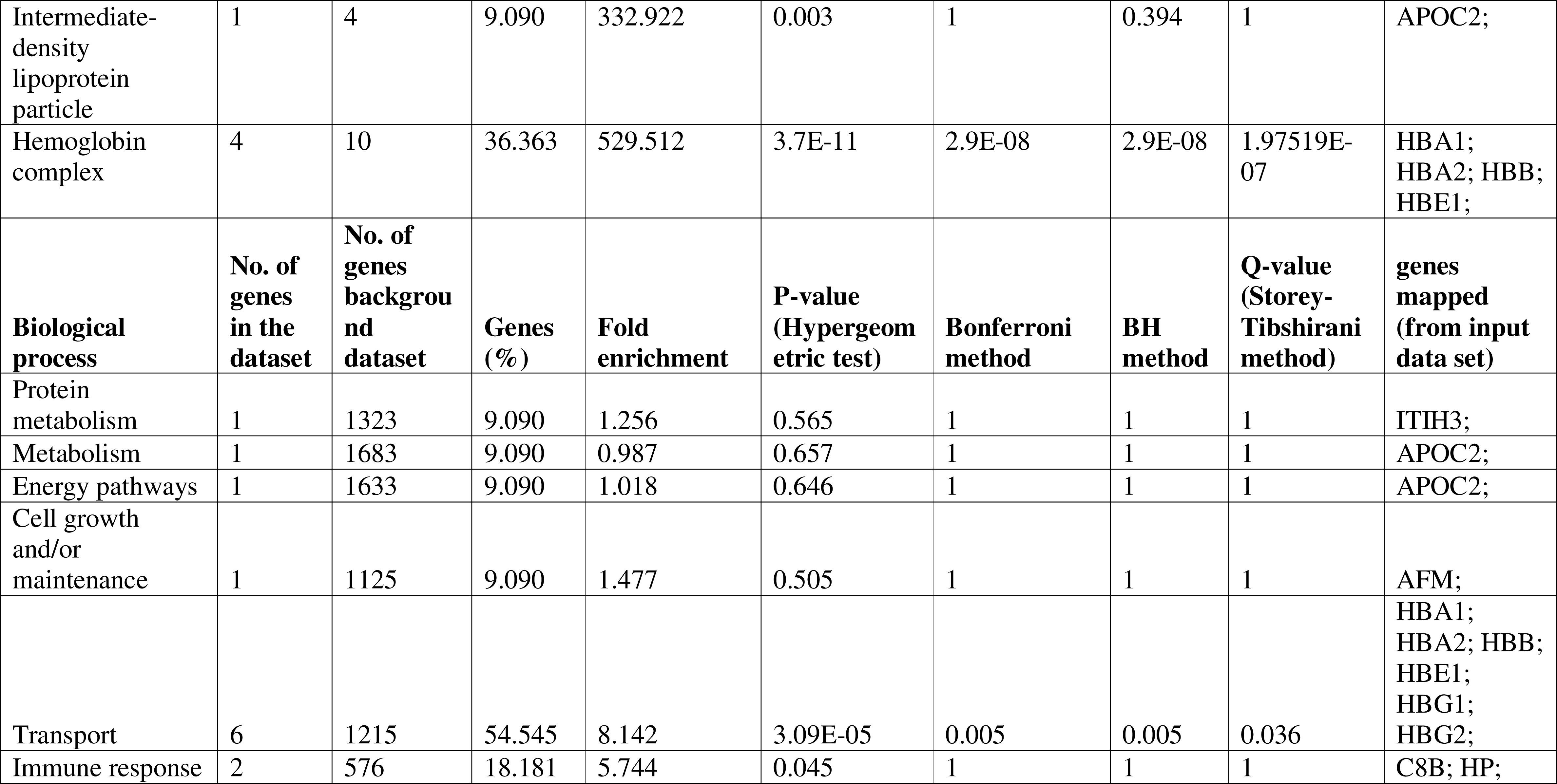
Biological process, cellular component, and molecular function of proteins found to be statistically significantly differentially expressed in the Amyotrophic Lateral Sclerosis (ALS) serum EVs compared to the HC serum EVs.

## Abbreviations

(ALS): Amyotrophic Lateral Sclerosis
(EVs): Extracellular Vesicles
(TDP-43): TAR DNA-binding protein 43
(SOD1): Superoxide Dismutase 1
(HC): Healthy control
(MVB): Multivesicular body
(BCA): Bicinchoninic acid
(SDS): Sodium dodecyl sulphate
(TFA): Trifluoroacetic acid
(ACN): Acetonitrile
(GO): Gene ontology
(MAC): Membrane attack complex
(CMA): Chaperone mediated autophagy
(Lamp2A): Lysosomal associated membrane protein 2A
(FTD): Frontotemporal Dementia

## Acknowledgments

This work was supported by the National Health and Medical Research Council (GNT1041413 to A.F.H.) and the MND Foundation, Australia (A.F.H.). N.V. is supported by an Australian Postgraduate Scholarship. We acknowledge the use of the La Trobe University Proteomics and Metabolomics Platform and the La Trobe University Imaging Facility for undertaking this research. Finally, we acknowledge P.L., E.B., A.F., and I.Q. for providing the samples.

## Author Contributions

L.C. and A.F.H. designed the study. P.L., E.B., A. F., and I.Q. provided the serum samples. N.V. processed the serum samples with help from J.S. N.V. performed characterization and prepared samples for mass spectrometry analysis. R.L. performed mass spectrometry analysis and interpreted the datasets and constructed the bioinformatic figures. N.V. performed the G.O. analysis. N.V. and L.C. constructed the manuscript.

## Data Availability Statement

Proteomics data are available via ProteomeXchange with identifier PXD036652.

